# Tooth Loss, Patient Characteristics, and Coronary Artery Calcification

**DOI:** 10.1101/2024.01.28.24301883

**Authors:** Tuan D. Pham, Lifong Zou, Mangala Patel, Simon B. Holmes, Paul Coulthard Barts, The London Faculty of Medicine and Dentistry

## Abstract

This study, for the first time, explores the integration of data science and machine learning for the classification and prediction of coronary artery calcium (CAC) scores, investigating both tooth loss and patient characteristics as key input features. By employing these advanced analytical techniques, we aim to enhance the accuracy of classifying CAC scores into tertiles and predicting their values. Our findings reveal that patient characteristics are particularly effective for tertile classification, while tooth loss provides more accurate predicted CAC scores. Moreover, the combination of patient characteristics and tooth loss demonstrates improved accuracy in identifying individuals at higher risk of cardiovascular issues related to CAC. This research contributes valuable insights into the relationship between oral health indicators, such as tooth loss, patient characteristics, and cardiovascular health, shedding light on their potential roles in predictive modeling and classification tasks for CAC scores.

## 1 Introduction

The intersection between oral health and cardiovascular well-being has been a subject of growing interest in the medical and dental research communities [1, 2, 3, 4, 5, 6, 7, 8, 9]. Among various oral health indicators, tooth loss has emerged as a potential marker linked to systemic health conditions, including cardiovascular diseases [10, 11].

In an early investigation, Holmlund et al. [12] presented a study with an extended follow-up period. Their aim was to explore whether various parameters of oral health exhibit an association with future mortality in distinct cardiovascular disorders. The finding highlighted a link between oral health and cardiovascular diseases, underscoring the suitability of the number of teeth as a meaningful indicator for oral health in this specific context.

Gao et al. [13] found that periodontitis stands as a risk factor for coronary heart disease (CHD), establishing a positive correlation between the number of extracted teeth and CHD risk. Recognizing the significance of these factors in clinical assessments is crucial due to their association with cardiovascular risks.

Additional insights from Cheng et al. [14] underscore a significant increase in the association between tooth loss and the risk of cardiovascular disease and stroke. Subgroup analyses revealed links, especially within Asian and Caucasian populations, and across both fatal and non-fatal cases. The study also identified a noteworthy dose-response relationship between tooth loss and the risk of cardiovascular disease and stroke.

Another study by De Angelis et al. [15] highlighted that individuals with over 18 missing teeth face a 2.5 times greater risk of cardiovascular disease, which exhibits associations with Type 2 diabetes mellitus, underweight, and obesity. These findings affirm a connection between cardiovascular disease and oral health.

Beukers et al. [16] contributed to the discourse by emphasizing tooth loss as an outcome of prevalent dental conditions, dental caries, and periodontitis, constituting 2% of the global burden of human diseases. The systematic review and meta-analyses conducted in this study demonstrated that a diminished number of teeth serves as a risk factor for atherosclerotic cardiovascular diseases and mortality.

Beyond tooth loss, patient characteristics play a pivotal role in cardiovascular risk assessment. Factors such as age, gender, body mass index (BMI), smoking habits, and comorbidities have been extensively studied in the context of cardiovascular health [17]. A meta-analysis by Wong et al. [18] highlighted the multifactorial nature of cardiovascular risk, emphasizing the need to consider a spectrum of patient characteristics for accurate risk assessment. Research has sought to elucidate the intricate connections between tooth loss, patient characteristics, and CAC [19].

In this study, a pioneering exploration is carried out, employing advanced data science of tensor decomposition [20] and generalized additive models (GAMs) [21] in machine learning to address the classification and prediction of CAC scores. In particular, the methods of tensor decomposition have been pervasively applied to many areas of life and medical sciences [22]-[31]. The focus centers on tooth loss and patient characteristics to elucidate the intricate interplay between these health indicators and the broader spectrum of cardiovascular well-being. Tensor decomposition, a powerful analytical tool, is designed to uncover multidimensional patterns within the dataset, offering an understanding of the intricate relationships that may influence CAC scores. Simultaneously, the application of GAMs provides a flexible and robust framework, capable of capturing non-linear relationships within the data. This flexibility is especially crucial in the context of medical data, where relationships between variables are often complex and multifaceted.

The primary objective of research endeavor is to investigate the precision of classifying CAC scores into tertiles and predicting their values. By leveraging the insights pro-vided by tooth loss and patient characteristics, this study seeks to unravel latent patterns and contribute to the evolving landscape of predictive modeling in cardiovascular health assessments.

## 2 Materials and Methods

### 2.1 Patient Data

This investigation utilized a publicly available dataset [19] comprising 212 patients gathered from three hospitals located in the Netherlands, of which 114 were male. The average age of the participants was 57.8 years, with a mean body mass index (BMI) of 28 kg/m^2^. Among the participants, 32 had diabetes, 85 experienced hypercholesterolemia, and 128 were undergoing treatment for hypertension. The smoking history for all patients was categorized into three groups: 86 were non-smokers, 86 were former smokers, and 39 were current smokers.

The inclusion criteria for this study encompassed patients for whom both a coronary artery calcium (CAC) score and an orthopantomogram (OPG) were available, with both assessments recorded within a maximum period of 365 days spanning from 2009 to 2017. The CAC score, determined using the Agatston method, was measured through computed tomography scans.

The count of present teeth included all visible teeth on the OPG, encompassing third molars and radices relictae. Pontics of fixed partial dentures and prosthetic dentures were excluded from the tally of present teeth. The number of missing teeth was calculated by subtracting the count of present teeth from the expected total of 32 teeth. Dental implants were individually recorded.

The patients’ CAC scores were divided into tertiles. The first tertile consists of the lowest CAC scores, the second tertile comprises intermediate CAC scores, and the third tertile includes the highest CAC scores.

### 2.2 Tensor decomposition of tooth loss and patient characteristics in coronary artery calcification

A tensor is a multiway or *n*-way array with different orders, where an order one tensor is a vector, an order two tensor is a matrix, and an order three tensor is a volume. An *n*-th order or *n*-way tensor takes the form of an *n*-hypershape.

In a general form, the elements of an *n*-way tensor, denoted as **<u>T</u>** (where the underline indicates a tensor), are given as [32, 33]

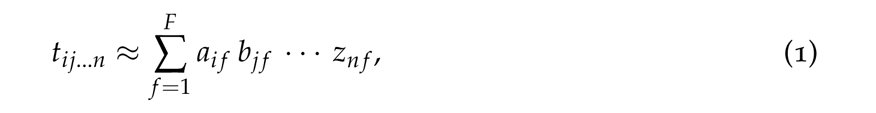

where *F* represents the number of factors, *t_ij_*_…*n*_ are the elements of **<u>T</u>**, and *a_i f_*, *i* = 1, . . ., *I*, *b_j f_*, *j* = 1, . . ., *J*, and *z_n f_*, *n* = 1, . . ., *N*, are elements of the loading matrices **A**, **B**, and **Z**, respectively.

Alternatively, a tensor can be expressed as

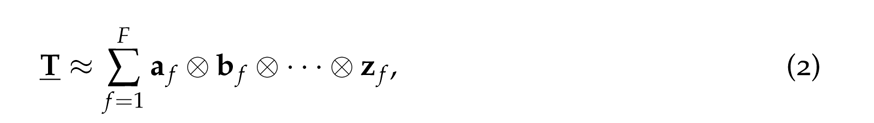

where *⊗* denotes the outer product, and **a** *_f_*, **b** *_f_*, and **z** *_f_* are the *f* -column vectors of **A**, **B**, and **Z**, respectively.

The loading matrices **A**, **B**, and **Z** can be computed using the PARAFAC decomposition model [34, 35].

To examine the separability of tooth loss in terms of different CAC tertiles for classifying tertiles and predicting coronary artery calcium scores, two-way tensors for three cohorts of patients with different CAC tertiles, denoted as **<u>T</u>***_k_*, can be modeled as:

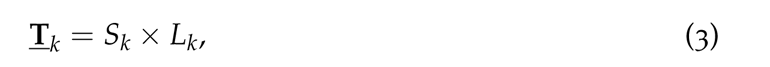

where *k* = 1, 2, 3 representing tertiles 1, 2, and 3, respectively; *S_k_* is the number of patients in tertile *k*, and *L_k_* is the corresponding numbers of missing teeth for patients in tertile *k*.

### 2.3 GAM for classification and prediction of coronary artery calcification

In statistical analysis, a GAM is a type of generalized linear model where the linear response variable is influenced by unknown smooth functions of specific predictor variables. This model establishes a connection between a univariate response variable, denoted as *y*, and a set of predictor variables, *x_i_*, *i* = 1, 2, . . ., *p*, which is mathematically expressed as

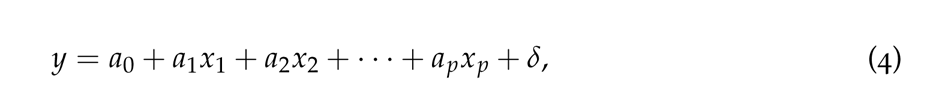

where *a*_0_, *a*_1_, *· · ·*, *a_p_* are estimated coefficients, and *δ* is the error term.

To accommodate non-linear effects, a GAM substitutes each linear component with a smooth function:

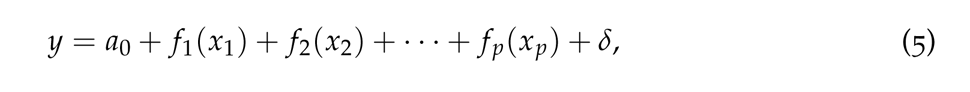

where *f_j_*, *j* = 1, 2, . . ., *p*, are smooth functions.

Equation (5) is referred to as a GAM since each smooth function is independently estimated, and the sum of these individual contributions is then combined. It is pointed out [36] that GAMs exhibit great flexibility because these models allow for distinct smooth functions corresponding to each predictor. Consequently, a GAM can incorporate various techniques, such as: a) non-linear polynomial methods for continuous predictors, b) step functions, particularly suitable for handling categorical predictors, and c) linear models, chosen when deemed more suitable for certain predictors.

In this study, GAM fitting was carried out by employing boosted trees [37] as smooth functions for the predictors. The fitting process involved constructing a set of predictor trees during each boosting iteration, with the initial learning rate determined through Bayesian optimization. In binary classification of CAC tertiles, the GAM yielded class scores (the logit of class probabilities) by summing univariate smooth functions of the predictors. In regression tasks for predicting CAC scores, the GAM generated a response variable through the aggregation of univariate smooth functions of the predictors.

### 2.4 Classification performance metrics

Let *P* represent the count of instances in higher CAC tertiles, *N* denote the count of instances in lower CAC tertiles, *TP* (true positive) indicate the count of correctly classified higher tertile cases, *TN* (true negative) be the count of correctly classified lower tertile cases, *FP* (false positive) stand for the count of misclassified higher tertile cases, and *FN* (false negative) refer to the count of misclassified lower tertile cases.

Table 1 defines the performance measures, including accuracy (*ACC*), sensitivity (*SEN*), specificity (*SPE*), precision (*PRE*), and *F*_1_ score. Additionally, the assessment of binary classification performance involves employing the receiver operating characteristic (ROC) curve. The ROC curve is constructed by plotting *TP* against *FP* across different thresholds. In certain contexts, the true positive rate (TP rate) corresponds to the probability of correct classification, while the false positive rate (FP rate) is the probability of false alarm. A higher area under the curve (AUC) in the ROC analysis indicates superior discriminative ability of the model.

**Table 1:** Performance measures of binary classification.

| $ACC$ | $SEN$ | $SPE$ | $PRE$ | $F_1$ score |
| --- | --- | --- | --- | --- |
| $\frac{TP + TN}{P + N}$ | $\frac{TP}{P}$ | $\frac{TN}{N}$ | $\frac{TP}{TP + FP}$ | $\frac{2TP}{2TP + FP + FN}$ |

## 3 Results

Figure 1 shows the 2D and 3D plots depicting three tensor-decomposition factors (*F*_1_, *F*_2_, and *F*_3_) obtained through the PARAFAC scheme. These factors were derived from the patient tooth-loss model for the three CAC tertiles.

**Figure 1:**
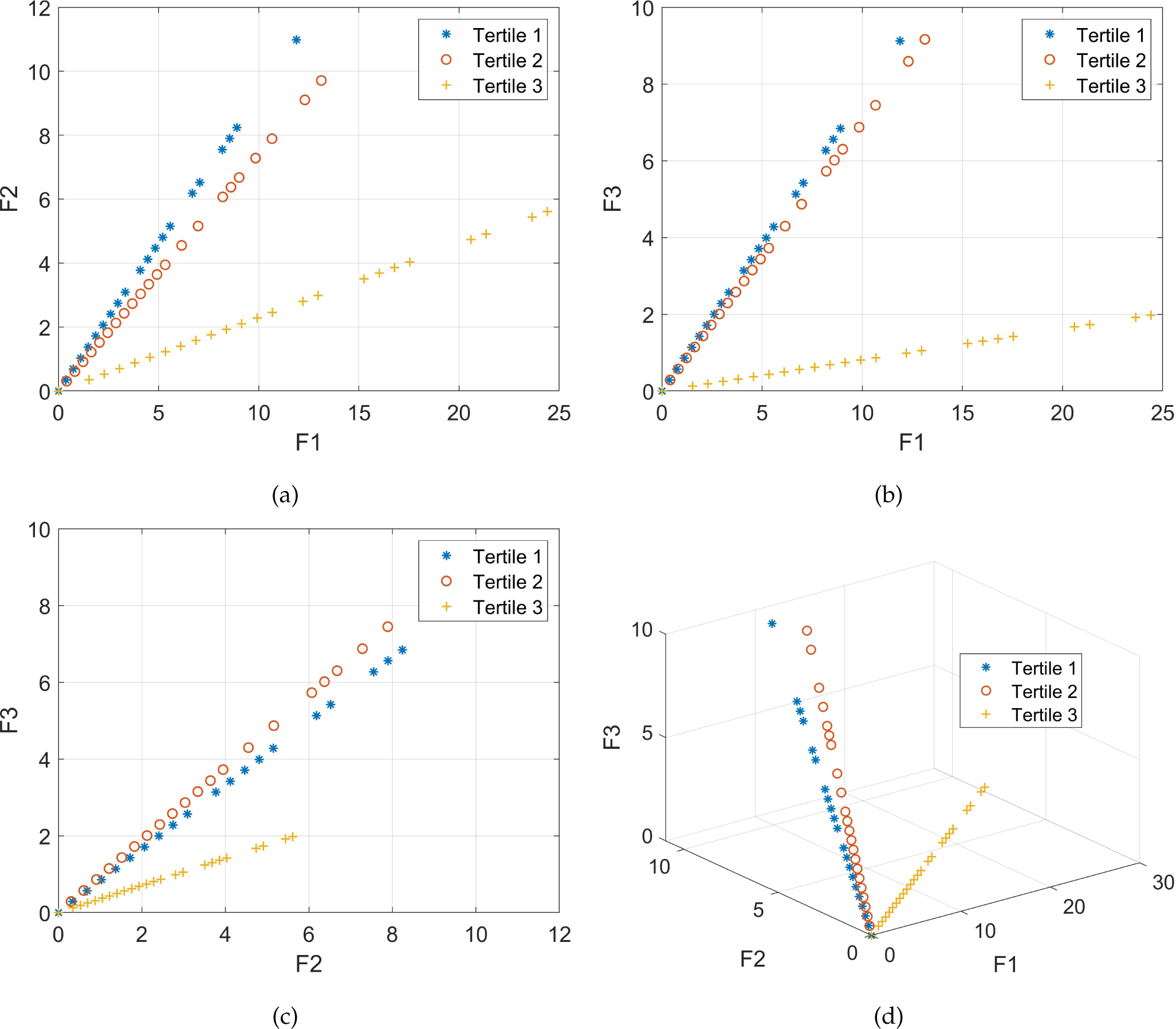
PARAFAC tensor-decomposition factors of patient tooth-loss model.

Table 2 shows the results obtained from a ten-fold cross-validation employing the univariate GAM for the classification of two distinct classes. In this context, class 1 includes the first tertile, whereas class 2 encompasses the second and third tertiles. The classification process used various input features, encompassing patient characteristics, tooth loss data, and the combination of patient characteristics with tooth loss information.

**Table 2:**
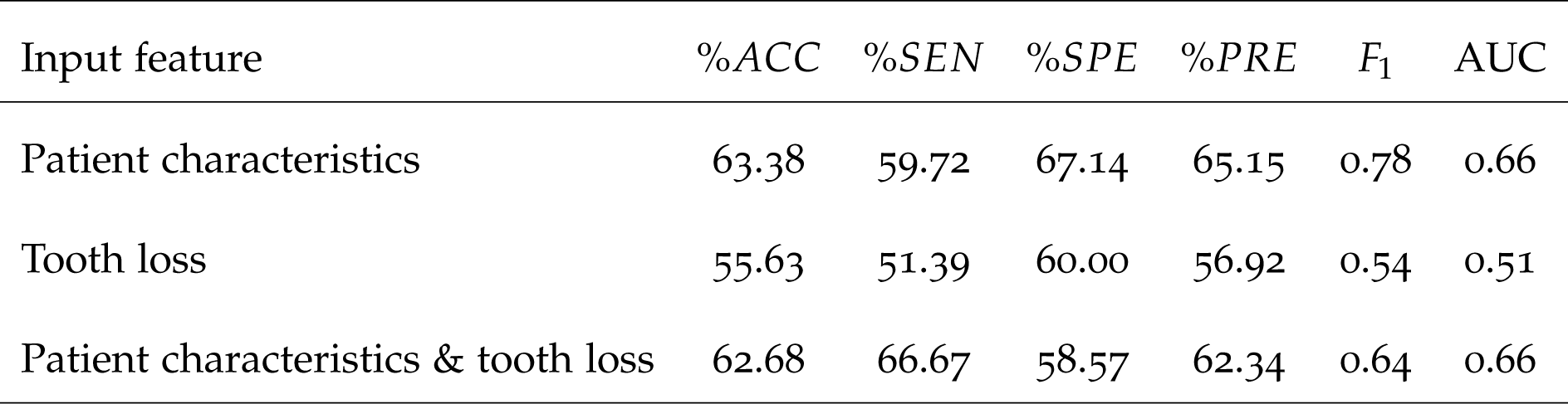
Ten-fold cross-validated GAM-based categorization of CAC into tertile 1 (class 1) and tertiles 2 & 3 (class 2).

Similarly, Table 3 exhibits the outcomes of a ten-fold cross-validation utilizing the univariate GAM for the classification of two distinct classes. In this instance, class 2 is defined by the second tertile, while class 3 is the third tertile. The classification process incorporated a diverse set of input features, including patient characteristics, tooth loss data, and the combination of patient characteristics with tooth loss information.

**Table 3:**
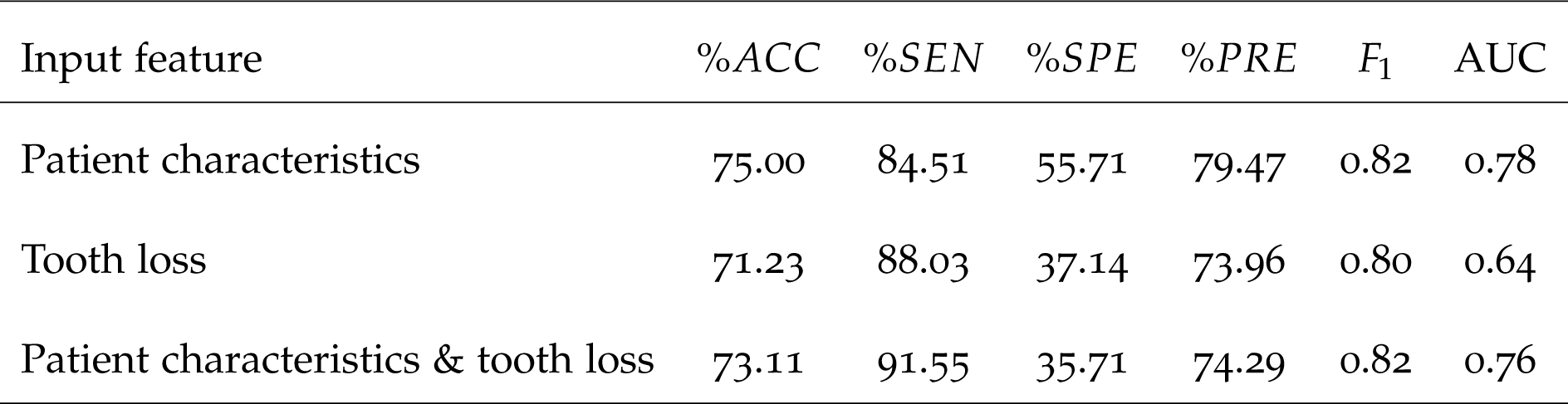
Ten-fold cross-validated GAM-based categorization of CAC into tertile 2 (class 1) and tertile 3 (class 2).

Figure 2 visually presents the confusion matrices and AUCs corresponding to the classification outcomes depicted in Tables 2 and 3.

**Figure 2:**
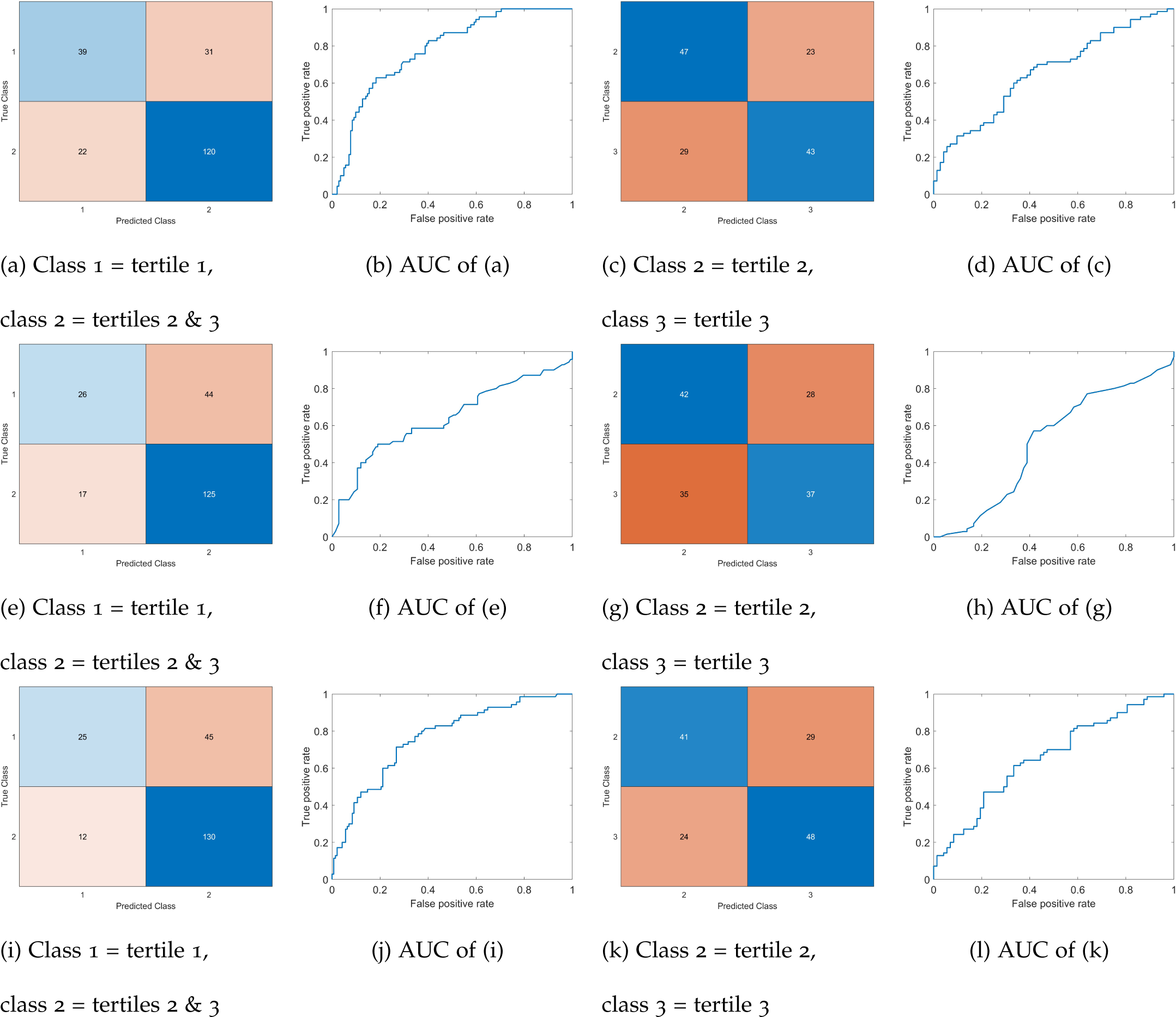
Confusion matrices and AUCs obtained from ten-fold cross-validated GAM-based binary classification of CAC tertiles using patient characteristics (a)-(d), tooth-loss (e)-(h), and combined patient characteristics and tooth-loss (i)-(l).

In the prediction analysis, a male patient with a CAC score of 20,000 was excluded from the dataset due to its singular and notably high value, which qualifies as an outlier. Table 4 shows the ten-fold cross-validation regression errors produced from the univariate GAM, using five input scenarios: patient characteristics, patient characteristics with tooth loss (both male and female), tooth loss (both male and female), female tooth loss, and male tooth loss. Figure 3 accompanies Table 4, visually portraying the predicted and observed CAC scores as derived from the univariate GAM for regression.

**Figure 3:**
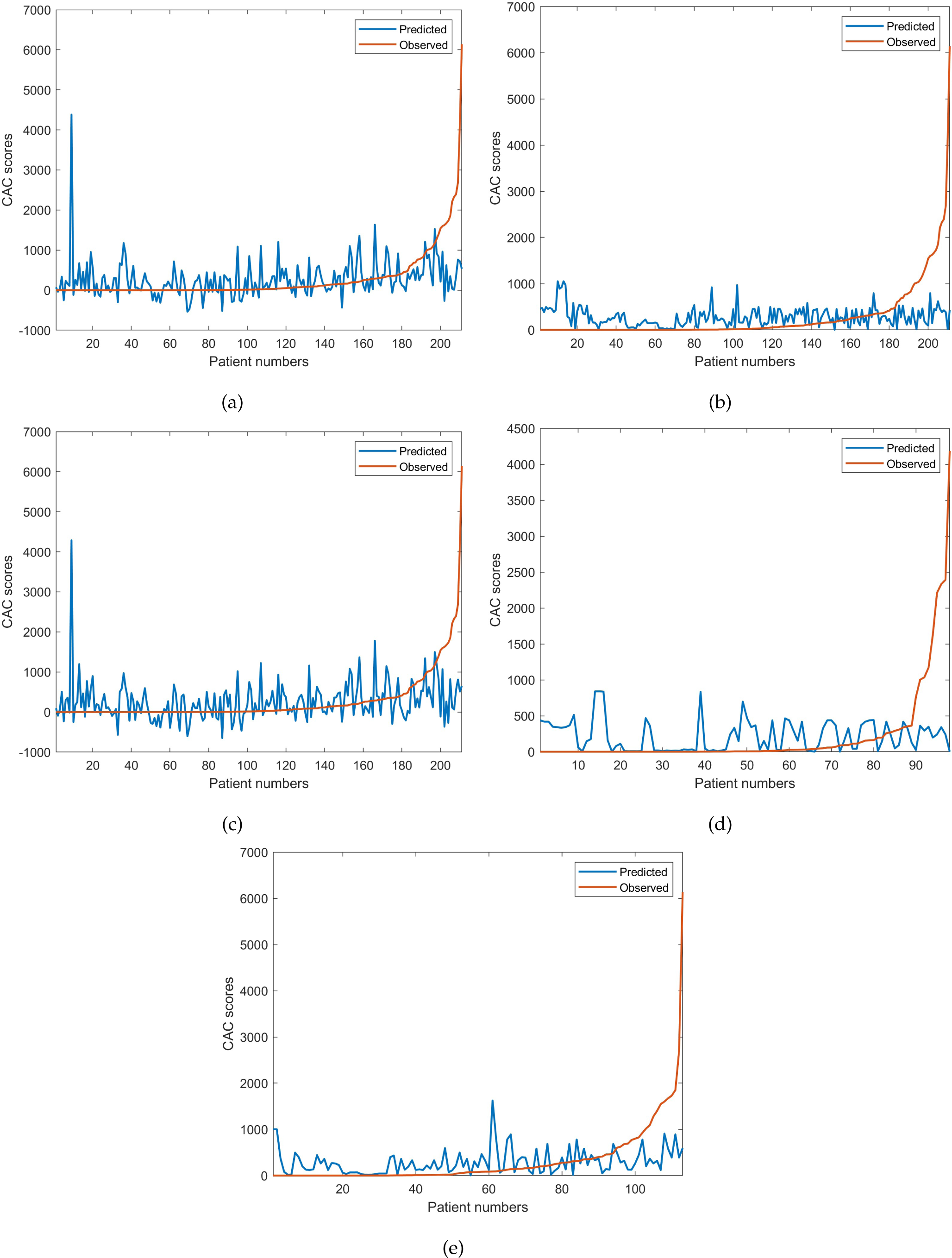
Observed CAC scores and predicted values obtained from ten-fold cross-validated GAM for regression using: (a) patient characteristics, (b) patient characteristics with male and female tooth loss, (c) male and female tooth loss, (d) female tooth loss, and

**Table 4:** Regression loss of ten-fold cross-validated GAM-based prediction of CAC scores.

| Input feature | Average mean squared error ($10^5$) |
| --- | --- |
| Patient characteristics | 5.816 |
| Tooth loss | 4.823 |
| Patient characteristics & tooth loss | 6.016 |
| Male tooth loss | 5.119 |
| Female tooth loss | 4.271 |

## 4 Discussion

Table 2 reveals insights into the classification of CAC tertiles, where the utilization of tooth loss as an input feature (71%) stands competitively against patient characteristics (75%) in terms of accuracy. Interestingly, the combination of patient characteristics and tooth loss (73%) does not yield an improvement in classification accuracy compared to using patient characteristics alone. It is also worth noting an observation: tooth loss, when employed as a singular feature, surpasses in identifying tertiles 2 and 3 (88%), which are indicative of a higher risk of coronary artery disease, outperforming the sensitivity achieved with patient characteristics alone (85%). However, the result obtained from the combined features of patient characteristics and tooth loss achieved the highest sensitivity (92%).

The AUC value (0.64) of using tooth loss, which are lower than the patient characteristics (0.78) and the combination of both features (0.76), as a singular input to the GAM suggest a more severe trade-off between sensitivity and specificity– indicating an imbalance between sensitivity and specificity, with the classifier favoring one at the expense of the other. Precision and *F*_1_ score obtained from the input of the patient characteristics (*PRE* = 79%, *F*_1_ = 0.82) are higher than those obtained from the tooth loss (*PRE* = 74%, *F*_1_ = 0.80) and the combined features (*PRE* = 74%, *F*_1_ = 0.82). However, the differences are minor between the three cases of input.

The above results underscore the intricate relationship between tooth loss, patient characteristics, and their independence in the classification of CAC tertiles. The observations also suggest that the combination of tooth loss and patient characteristics information can enhance the ability to detect higher-risk categories, emphasizing the importance of considering both dental and patient-specific factors in refining the sensitivity of CAC tertile classification.

The 2D and 3D plots depicting the three PARAFAC tensor-decomposition factors de-rived from the tooth-loss model becomes evident in their ability to effectively distinguish between the three CAC tertiles. This observation holds meaningful implications, suggesting that tooth loss can serve as a useful feature for the classification and prediction of CAC scores. The separability demonstrated in the graphical representations underscores the potential utility of tooth loss as a valuable marker in understanding and predicting coronary artery calcium levels.

The classification between tertiles 2 and 3, as illustrated in Table 3, reveals a discernible drop in performance compared to the differentiation of tertile 1 from both tertiles 2 and 3. The performance metrics derived from the patient characteristics feature and the combination of patient characteristics and tooth loss exhibit similarities, while the metrics stemming from tooth loss alone register a lower performance.

Examining the specificities, the combined features of patient characteristics and tooth loss demonstrate enhanced capability in identifying CAC tertile 3 (67%), which is indicative of higher risk of coronary artery disease. On the other hand, patient characteristics alone exhibit superior performance in pinpointing CAC tertile 2 (67%). These differences in performance underscore the differential contributions of individual and combined features in classifying CAC tertiles, shedding light on the potential of specific feature combinations for improved predictive modeling in coronary health assessment.

To gain a deeper understanding of the classification of CAC tertiles, Figures 4 and 5 illustrate the local and partial dependence effects [38], showcasing examples of correctly and incorrectly classified CAC tertiles 2 and 3 in both male and female subjects.

**Figure 4:**
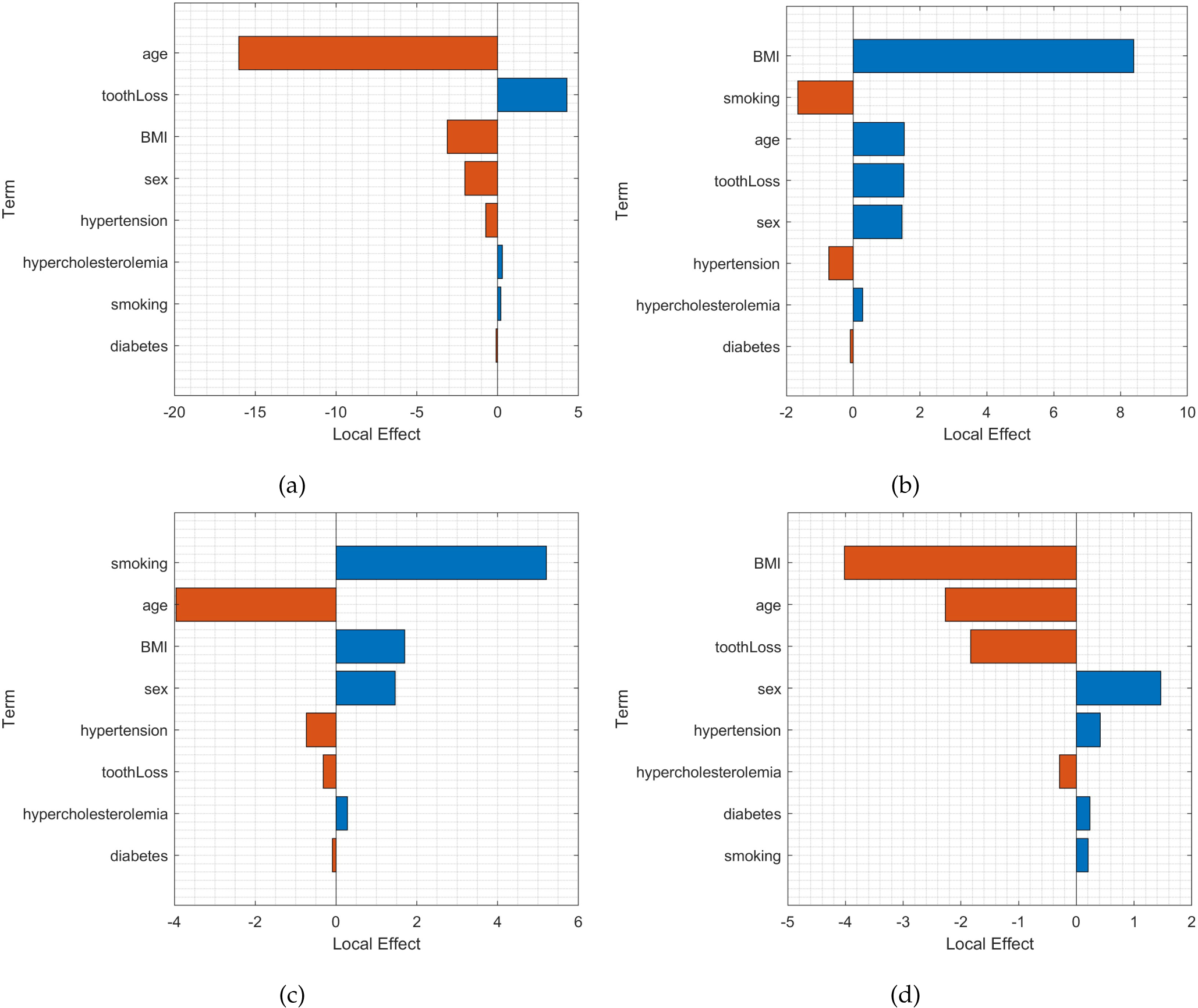
Local effects in: (a) correct classification of tertile 2 for a female patient (age = 36 years, smoking = never, diabetes = no, hypercholesterolemia = no, hypertension = no, BMI = 30.3, and tooth loss = 12); (b) correct classification of tertile 3 for a male patient (age = 66 years, smoking = never, diabetes = no, hypercholesterolemia = no, hypertension = no, BMI = 22.64, and tooth loss = 32); (c) misclassifying tertile 2 as tertile 3 for a male patient (age = 73 years, smoking = never, diabetes = yes, hypercholesterolemia = yes, hypertension = yes, BMI = 25.25, and tooth loss = 7); and (d) misclassifying tertile 3 as tertile 2 for a male patient (age = 56 years, smoking = never, diabetes = no, hypercholesterolemia = no, hypertension = no, BMI = 23.99, and tooth loss = 16).

**Figure 5:**
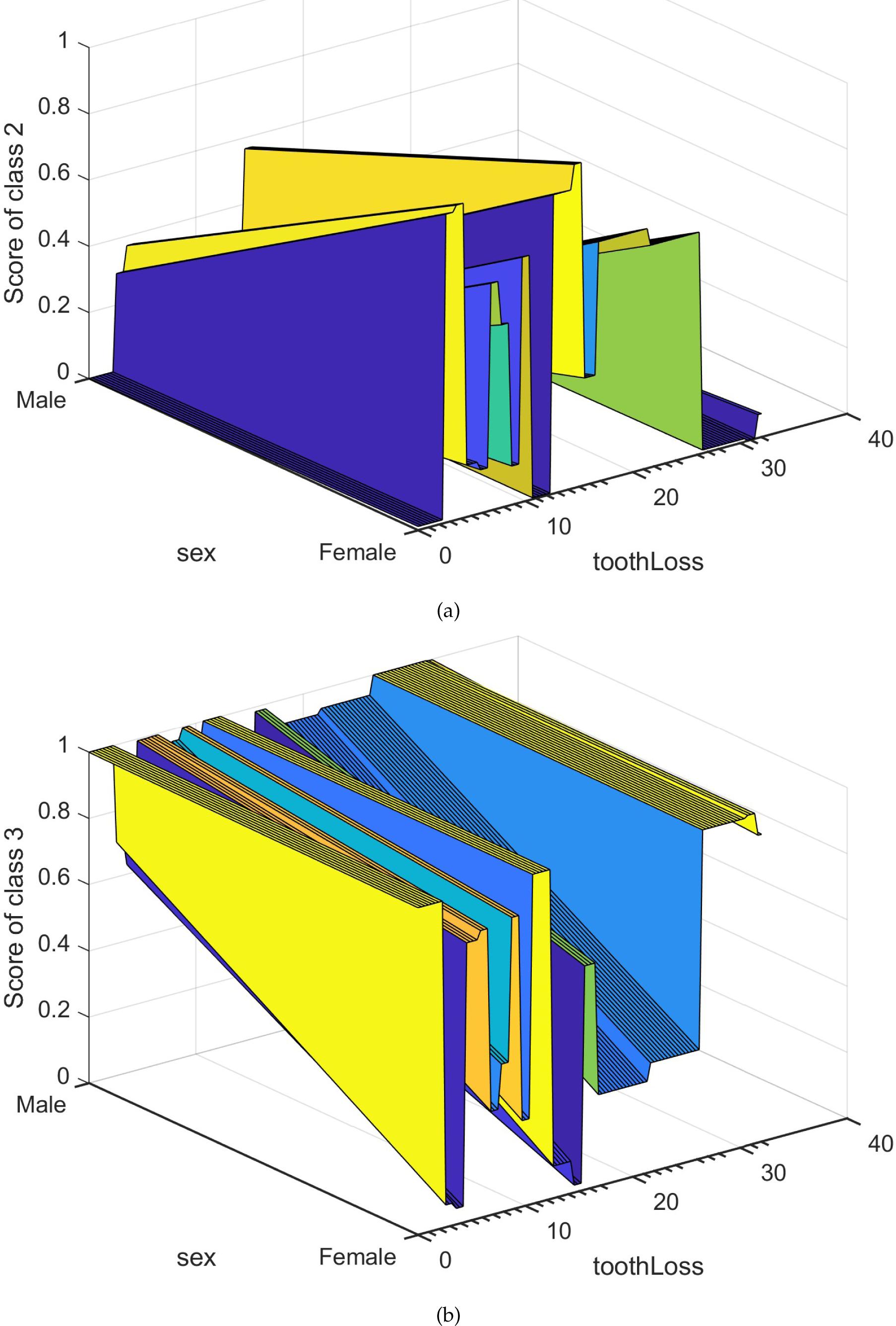
Partial effects in classifying: (a) class 2 (tertile 2), and (b) class 3 (tertile 3) in terms of tooth loss and sex.

In Figure 4, the bar graphs represent the local effects of clinical and tooth loss attributes on GAM-based classification. Each local effect value denotes the contribution of a specific term to the classification score for a given tertile, which is the logit of the posterior probability for that tertile in the observation.

For the accurate classification of tertile 2 in a female patient (Figure 4 (a)), tooth loss emerges as the most influential factor. In the correct classification of tertile 3 in a male patient (Figure 4 (b)), BMI takes precedence, while tooth loss, age, and sex contribute equally to a positive influence. On the other hand, the misclassification of tertile 2 as tertile 3 in a male patient (Figure 4 (c)) is driven by smoking, BMI, and sex. Similarly, the misclassification of tertile 3 as tertile 2 in a male patient (Figure 4 (d)) is primarily influenced by sex and hypertension.

Moving to the partial dependence effects in Figure 5 (a) and (b), these illustrate the partial dependence of score values for classifying tertile 2 (class 2) and tertile 3 (class 3) based on sex and tooth loss, respectively. The plots suggest that tooth loss has a more pronounced impact on the classification of tertile 2 in female patients, while tooth loss in male patients exerts a stronger influence on the classification of tertile 3.

Concerning the CAC score prediction utilizing GAM for regression, as depicted in Table 4 and Figure 3, the inclusion of tooth loss as an independent feature yielded the smallest average mean squared error (4.82 *×* 10^5^). Conversely, the integration of patient characteristics and tooth loss resulted in the highest error (6.02 *×* 10^5^), with patient characteristics alone falling in between the other two input features (5.82 *×* 10^5^).

Regarding the prediction of male and female CAC scores, both use of patient characteristics and the combined input of patient characteristics and tooth loss resulted in largest errors. The use of individual tooth loss feature is more favorable, where the female tooth loss resulted in the smallest error. As shown in Figure 3, large errors resulted in the regression model were mainly due to some relatively very large observed CAC scores of the patients indexed toward the right end of the plots.

In the context of CAC score prediction using GAM for regression, as shown in Table 4 and Figure 3, the incorporation of tooth loss as an independent feature led to the lowest average mean squared error (4.82 *×* 10^5^). Conversely, when patient characteristics and tooth loss were combined, the highest error was observed (6.02 *×* 10^5^), while using patient characteristics alone fell in between the errors associated with the other two input features (5.82 *×* 10^5^). Concerning the prediction of male and female CAC scores, opting for the individual tooth loss feature, female tooth loss yielded the smallest error (4.27 *×* 10^5^).

As depicted in Figure 3, substantial errors in the regression model primarily arose from some exceptionally large observed CAC scores of patients situated towards the right end of the plots.

The occurrence of significant errors can be attributed to the imbalance in the dataset, leading to bias in the regression learning process. This imbalance stems from the disproportionate distribution of very small and very large CAC scores, with the majority of values falling within the category of very small scores. The dataset exhibits a skew toward these lower values, contributing to the observed imbalance.

## 5 Conclusion

The exploration of patient characteristics and tooth loss information in the classification and prediction of CAC scores has been presented and discussed in the preceding sections. The results suggest that patient characteristics is advantageous in the classification of tertiles, while tooth loss exhibits the potential for providing more accurate predicted CAC scores.

With the availability of additional data that addresses the bias in small CAC scores, there is a strong anticipation that the accuracies of GAM-based score prediction and tertile classification will significantly improve. The integration of patient characteristics and tooth loss emerges as a promising avenue, contributing to enhanced accuracy in identifying individuals at a higher risk of cardiovascular health issues, particularly in the realm of coronary artery calcium.

## Data Availability

This public dataset can be freely downloaded at: https://figshare.com/articles/dataset/S1_Data_-/13391239

https://figshare.com/articles/dataset/S1_Data_-/13391239

